# Use of Probiotics in the Prevention of Clostridioides difficile Infections during Antibiotic Exposure: A Systematic Review and Meta-Analysis

**DOI:** 10.1101/2023.10.04.23296557

**Authors:** Veronica Dabrowski

**Affiliations:** School of Medicine, University of Limerick, Ireland; University of Limerick, Limerick, Ireland, V94 T9PX

**Keywords:** probiotics, *Clostridioides difficile infection*, diarrhea, antibiotics, meta-analysis

## Abstract

*Clostridioides difficile* infections are a growing concern in the hospital setting. Current prevention methods include infection control strategies, antibiotic stewardship, and proper hand hygiene. However, the occurrence of *C. difficile* still manages to cause nosocomial outbreaks especially in vulnerable populations. Probiotics have been historically questioned for their use in the prevention of antibiotic-associated diarrhea and more specifically, *C. difficile* infections.

This meta-analysis pools 10 randomized controlled trials for the prevention of *Clostridioides difficile* infections (CDI) from reviewing the Cochrane Central Register of Controlled Trials (CENTRAL). *A priori* inclusion criteria remained as follows: RCTs, blinded/open trials, all populations, articles, or conference abstracts. Exclusion criteria excluded publications published outside 2013-2023*, non-English language trials, pre-clinical trials/protocols, case reports/series/systematic reviews, duplicates, probiotics not specified in methods, non-RCTs, incomplete/no outcomes reported, no confirmation of *Clostridioides difficile* infection. Probiotic strains tested in these trials mainly included *Lactobacillus spp. and Bifidobacterium spp*. Some studies showed significant benefits while others did not find significant improvement in the prevention of *C. difficile* infections.

The meta-analysis suggests that probiotics may have a positive effect in preventing CDI during antibiotic treatment. The study results had large levels of statistical heterogeneity which indicates an argument for further large-scale research to provide more definitive evidence on the efficacy of probiotics in CDI prevention.

## 1. Introduction

*Clostridioides difficile* is a gram-positive anaerobic, spore and toxin-forming bacterium that is commonly found in the large intestine (1). It was originally termed, “difficult Clostridioides” due to the difficulty in culturing and isolating its growth on media (1). A proportion of the general population carries a small amount of *C. difficile* within their gut and do not experience any clinical symptoms. The gut microbiome, a mosaic of bacteria, fungi, and viruses that live within our gut, has been considered as a part of our innate immune system (2). The gut’s microbiota is suspected in keeping *C. difficile* pathogen levels to a tolerable level before experiencing clinical symptoms (1). However, the administration of antibiotics may cause “dysbiosis”. Dysbiosis refers to the alteration in the gut microbiota within the large intestines and allows for the development of antibiotic-associated diarrhea (AAD) (2,3). *C. difficile* is the primary cause of AAD(1). Clinical symptoms of *Clostridioides difficile* infection (CDI) include: loose stools, abdominal pain and cramping, fever (low-grade), nausea, and anorexia (1).

In 2021, there were a total of 1,542 new CDI cases reported to the Health Protection Surveillance Centre (HPSC) in Ireland (4). The mean age at time of reporting was 68.1 years with the highest proportion of those infected over the age of 65 (4). It is noted that over half of all reported cases are associated with a healthcare setting. 3% of reported cases (*n* = 35) resulted in severe CDI which includes either intensive care unit admission, colectomy surgery, or death (4). The most associated risk factors for CDI include antibiotic use, advanced age, and gastric acid suppression. Antibiotics most commonly associated with CDI include: clindamycin, broad-spectrum penicillins and cephalosporins, and fluroquinolones (1). Advanced age (defined as >65 years of age) is associated with increased severity and frequency of *C. difficile* infections. In a 2002 CDI outbreak in Quebec, Canada, the frequency of infection was 10-fold higher than that observed in younger adults (5). Although a causal association between CDI and gastric acid suppressor use has not been established, several studies have shown that gastric acid suppression is associated with increased risk of *C. difficile* infection.

Current prevention strategies for *C. difficile* infections in Ireland include antibiotic stewardship, avoiding gastric acid suppression, proper infection prevention and control strategies in healthcare setting, and adaptation of the SIGHT Mnemonic UK protocol (Suspect, Isolate, Gloves, Hand Hygiene, and Test) (6). Due to the continuous nosocomial outbreaks of CDI, innovative prevention strategies, such as the use of probiotics, are being studied (3,7). Probiotics are referred to as “good bacteria” or bacteria that live currently within the large intestine and make up the gut microbiota, which is vital to several biological processes.

This systematic review hopes to focus solely on the use of probiotics on the prevention of *Clostridioides difficile* infections during periods of antibiotic use. The use of meta-analysis enables us to pool data from several studies to assess general probiotic use in the hospital setting for the prevention of CDI.

## 2. Methods

### 2.1. Aims

The aim of this review is to assess the use of probiotics for prevention of *C. difficile* infections. Prevention of CDI is defined as individuals without diarrheal symptoms who are exposed to antibiotics and are given the intervention arm who do not develop diarrhea associated with a positive assay for *Clostridioides difficile*.

### 2.2. Search Strategy

This systematic review adhered to PRISMA (Preferred Reporting Items for Systematic reviews and Meta-Analysis) statement guidelines (8) as well as *a priori* inclusion and exclusion criteria. Due to the scope of a systematic review only the Cochrane Central Register of Controlled Trials (CENTRAL) (2013-2023) was searched. Search terms included: probiotics, *Clostridioides difficile*, and antibiotic associated diarrhea. In addition, Boolean operators and other relevant synonyms were used to ensure comprehensive coverage of the literature. Search strategy on the Cochrane Database was broad initially and then the .csv file was evaluated to narrow to the language, year of publication, and study type of interest (random controlled trials). Abstracts of the retrieved studies were reviewed and highlighted for inclusion. Full articles, if available, were retrieved as free full texts. If they were not readily available, a login to the University of Limerick’s Glucksman Library was performed to achieve maximum access to journal and article availability.

### 2.3. Inclusion and Exclusion Criteria

Inclusion criteria included randomized control trials (RCTs), blinded or open trials, adult/ pediatric/elderly populations whether inpatient or outpatient, articles or conference abstracts were accepted if full data was available.

Exclusion criteria included articles published outside of the year reference range (2013-2023)*\**, non-English language trials, pre-clinical studies or study protocols, case reports or case series, duplicate reports, probiotic strains not specified, non-randomized controlled trials, incomplete or no outcomes reported, or no assays for *Clostridioides difficile* were all excluded.

*\*Until July 23, 2023*.

### 2.4. Data Extraction

The data for meta-analysis was manually extracted from the 10 articles included in this review. Abstracts in conference journals that did not provide sufficient data were either excluded or PubMED/EMBASE/Cochrane were searched for a fully published article by the authors in question. Using a standardized data extraction spreadsheet, the following data was extracted: authors, journal, year of publication, population data, study aims, intervention data, randomization, degree of blinding, type of *C. difficile* assays performed, results, adverse effects, discussion, and registration of clinical trial.

### 2.5. Interventions

The trials included had participants who were randomized to either an intervention arm or control arm. The control groups all had a placebo (blinded). All types of probiotic formulation (eg., capsule, tablet, drink, etc.) were included in this review. All strains of probiotics were also included whether they were mono-species or multi-species interventions.

### 2.6. Statistical Analysis

Statistical analysis was performed using MAVIS v1.1.3 software. MAVIS’s computational back-end packages include R version 4.3.0, R Studio, compute.es 0.2.5, ggplot2 3.4.2, Mac 1.1.1, Mad 0.8.3, metafor 4.0.0, quantreg 5.95, SCM 1.3.1, SCRT 1.3.1, SCRT 1.3.1, irr 0.84.1, and weightr 2.0.2. Packages used for the graphical user interface include shiny 1.74, shinyAce 0.4.2, and shinyBS 0.61.1. Full information on MAVIS v1.1.3 can be found here: http://kylehamilton.net/shiny/MAVIS/(9). Meta-analysis was conducted for the CDI primary outcomes to calculate pooled relative risk and 95% confidence intervals using the Laird method.

Heterogeneity was assessed using effect size and sampling variance and applied via the random-effects model. If the studies were homozygous the fixed-effects model would be applied, but it heterozygous the random-effects model would be used. If significant heterogeneity was determined to be present, a sub-group analysis would be performed. To determine sources of heterogeneity, sub-group analysis would be performed using *a priori* sub-groups such as adults *versus* pediatrics, low-dose probiotics *versus* high-dose probiotics and lastly small number of probiotic strains *versus* large number of probiotic strains. The between study variance was assessed using a sub-group co-variate of tau^2^ estimates.

### 2.7. Publication Bias

Publication bias was assessed using the funnel plot approach and the fail-safe N calculation using the Rosenthal approach. In the absence of publication bias, the funnel plot approach resembles a symmetrically inverted funnel. The Fail Safe N Calculation estimates how many additional studies with non-significant or null results are required to be added into the meta-analysis to make the overall results non-significant.

## 3. Results

### 3.1. Initial Screening of Data Search

**Figure 1** provides a PRISMA diagram breakdown of the research screening process. The literature initially yielded 193 abstracts relating to probiotics and *Clostridioides difficile* that were screened for inclusion. Of the 193 records, 91 were removed before screening: year of publishing did not occur in 2013-2023 (n = 81), paper was in non-English language (n = 8), and paper was designated as a systematic review (n = 2). Next a .csv file was downloaded which contained n = 102 abstracts to be reviewed. After review, a total of 70 records were excluded: pre-clinical published abstract (n = 33), not *C. difficile* specific (n = 10), no probiotic was mentioned (n = 9), duplicate abstract (n =8), study type other than RCT (n = 7), no placebo arm (n = 2), and no article associated with abstract (n = 1).

**Figure 1.**
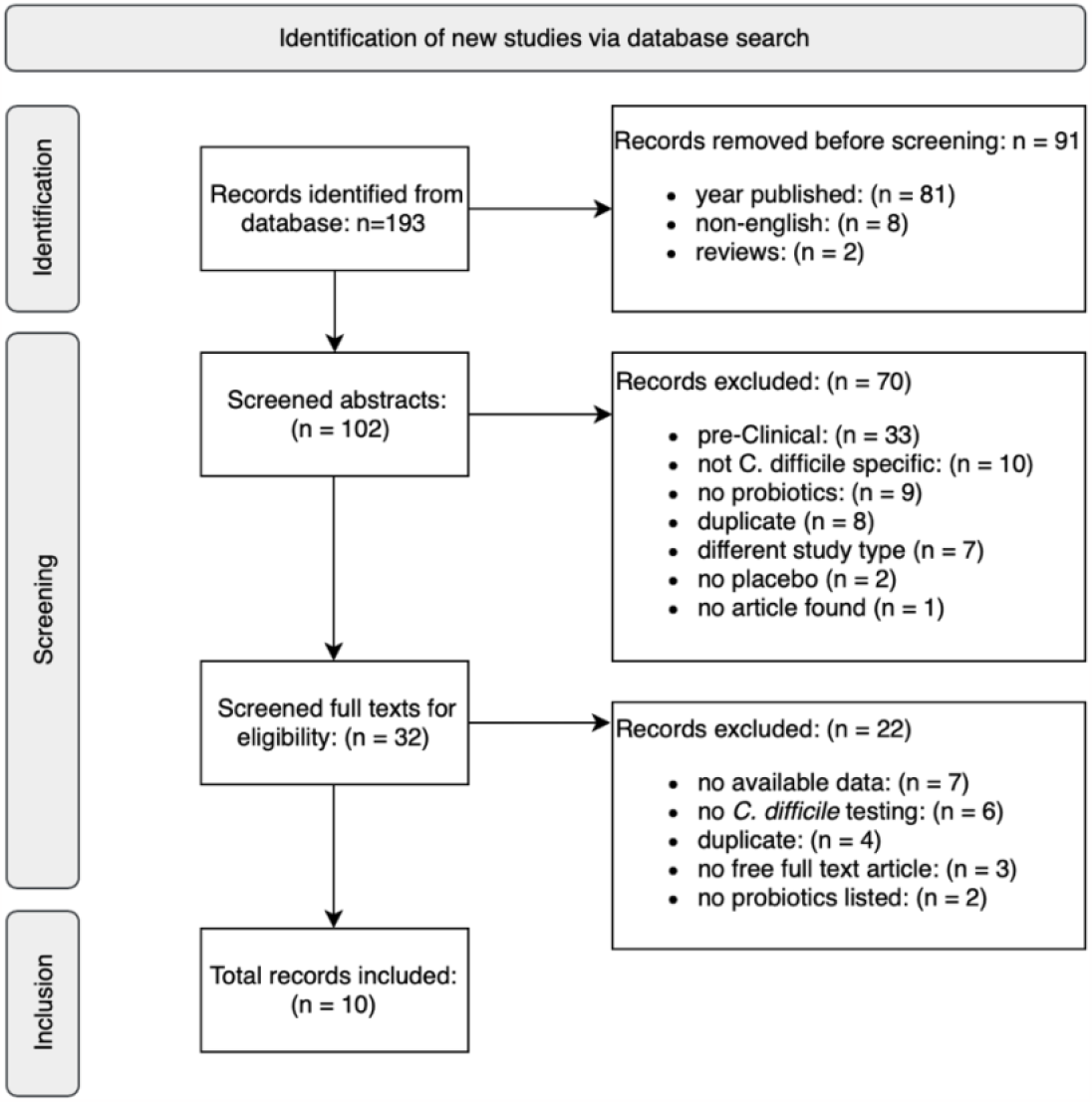
PRISMA flowchart of the literature search strategy and results performed for the use of probiotics for the prevention of *Clostridioides difficile infections*.

### 3.2. Secondary Screening of Full Articles

Of 102 screened abstracts, 32 full text articles were further screened with a total of 22 being excluded: no data was available (n = 7), no *C. difficile* testing performed (n = 6), duplicate article of an abstract (n = 4), no free full text article was available (n = 3), and no probiotic strains were listed (n = 2). A total of 10 records were included in the systematic review and meta-analysis, 3 of which were abstracts and the remainder were journal articles.

### 3.3. Included Trials

For the prevention of *Clostridioides difficile* infection, a total of 10 studies were included. Seven of which were journal articles and three of which were conference abstracts. For the 10 clinical trials, there were a total of 14 treatment arms which were included (10–19). Most of the studies included were from peer-reviewed, full text articles (*n*= 7, 70%), but three (*n* = 3, 30%) were only available as meeting abstracts. **Table 1** shows a breakdown of the trial populations. Population sizes of the random controlled trials ranged from 32 to 2,941 participants. A total of 2,743 participants were in the treatment arm and 2,403 participants were in the placebo arm. All the articles included in the systematic review and meta-analysis were in English only as this was part of the initial *a priori* inclusion and exclusion criteria prior to screening. The ten random controlled trials were conducted in seven different countries: United Kingdom (*n* = 4, 40%), United States of America (*n* = 1, 10%), Canada (*n* = 1, 10%), Bulgaria (*n* = 1, 10%), China (*n* = 1, 10%), Poland (*n* = 1,10%), and Japan (*n* = 1, 10%). The trials included a variety of hospitalized patients with *n* = 4 trials (40%) strictly recruiting patients that were hospitalized and would require acute antibiotic treatment, spinal cord injury patients accounted for 20% of the studies (*n* = 2), one trial included patients for elective colon cancer surgery (10%), one trial included patients specifically with respiratory infections, urinary tract infections and surgery prophylaxis (10%), one trial included strictly intensive care unit patients (10%), and one trial included gastroenterology patients only (10%). The trials were carried out in 15 hospital or specialized centres/departments.

**Table 1.**
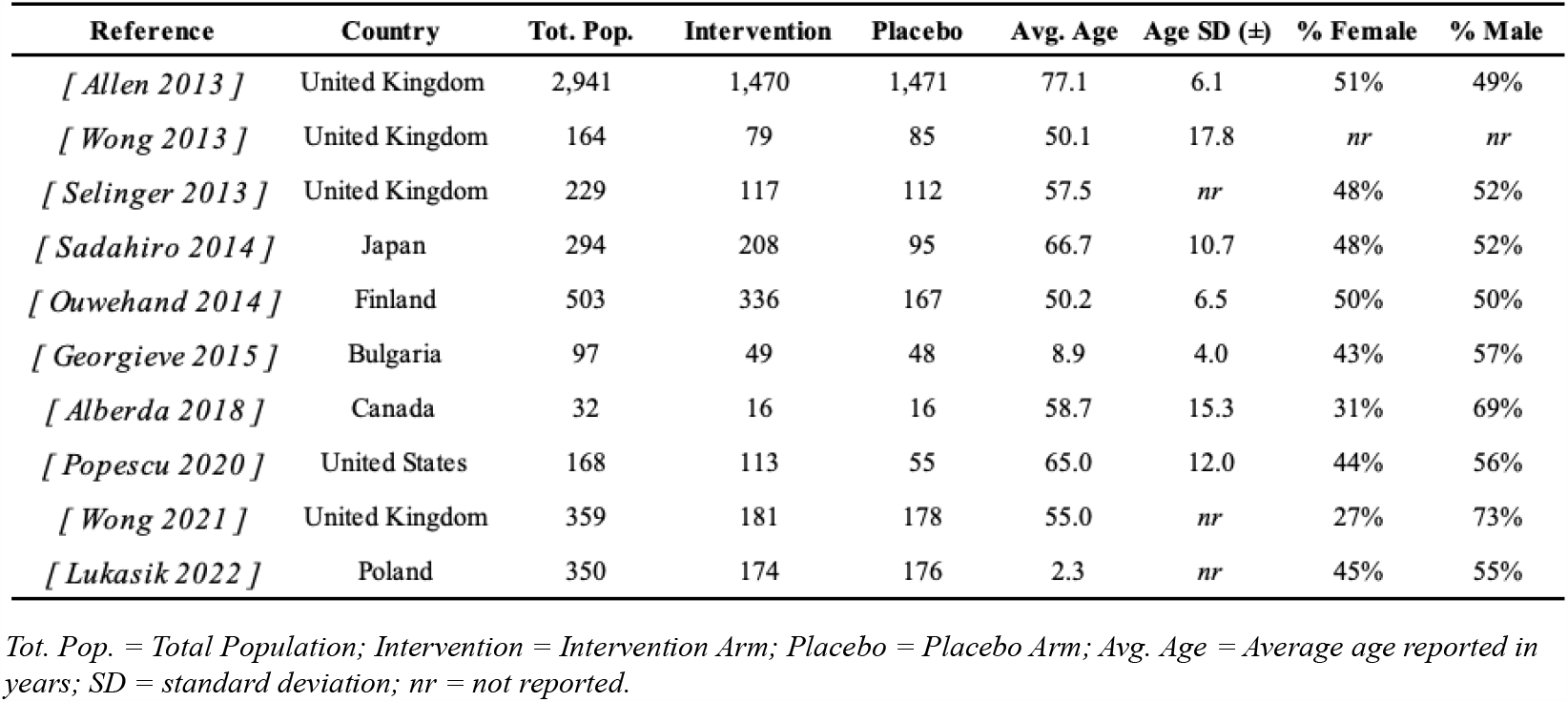
Population data from the 10 trials included in the systematic review and meta-analysis.

| Reference | Country | Tot. Pop. | Intervention | Placebo | Avg. Age | Age SD (±) | % Female | % Male |
| --- | --- | --- | --- | --- | --- | --- | --- | --- |
| [ Allen 2013 ] | United Kingdom | 2,941 | 1,470 | 1,471 | 77.1 | 6.1 | 51% | 49% |
| [ Wong 2013 ] | United Kingdom | 164 | 79 | 85 | 50.1 | 17.8 | *nr* | *nr* |
| [ Selinger 2013 ] | United Kingdom | 229 | 117 | 112 | 57.5 | *nr* | 48% | 52% |
| [ Sadahiro 2014 ] | Japan | 294 | 208 | 95 | 66.7 | 10.7 | 48% | 52% |
| [ Ouwehand 2014 ] | Finland | 503 | 336 | 167 | 50.2 | 6.5 | 50% | 50% |
| [ Georgieve 2015 ] | Bulgaria | 97 | 49 | 48 | 8.9 | 4.0 | 43% | 57% |
| [ Alberda 2018 ] | Canada | 32 | 16 | 16 | 58.7 | 15.3 | 31% | 69% |
| [ Popescu 2020 ] | United States | 168 | 113 | 55 | 65.0 | 12.0 | 44% | 56% |
| [ Wong 2021 ] | United Kingdom | 359 | 181 | 178 | 55.0 | *nr* | 27% | 73% |
| [ Lukasik 2022 ] | Poland | 350 | 174 | 176 | 2.3 | *nr* | 45% | 55% |

*Tot. Pop.* = Total Population; *Intervention* = Intervention Arm; *Placebo* = Placebo Arm; *Avg. Age* = Average age reported in years; *SD* = standard deviation; *nr* = not reported.

### 3.4. Study Design

#### 3.4.1. Degree of Blinding

Most of the random controlled trials (*n* = 7, 70%) were double-blinded, meaning that the placebo group was administered a product that was identical in appearance to that of the probiotic intervention group. One of the trials was listed as a “triple-blind” study (*n* = 1, 10%), where not only are the participants and the researchers blinded to the intervention and the placebo arms, but also the data analyzers (14). Two of the studies (20%) were unblinded. One of which contained intensive care unit patients (16) and therefore these patients are not aware of what is being administered to them and the second study included patients that were admitted to the gastroenterology department (17).

#### 3.4.2. Attrition in Prevention Trials

Attrition percentages for the trials ranged from 0%-47%, with two studies not reporting their attrition rates, likely since these were conference abstracts (10,17). Results can be viewed in **Table 2**. Of the studies that reported their attrition rates, one reported that they had zero attrition (*n* = 1, 12.5%). Two studies reported that they had low attrition rates (1%-10%) (*n* = 2, 25%), another 3 studies reported moderate attrition rates at 11%-20% (*n* = 3, 37.5%), one study reported high attrition rates 21%-40% (*n* = 1, 12.5%), and one study reported very high attrition rate from 41%-60% (*n* = 12.5%).

**Table 2.** Attrition rates and number of sites for each trial included in the meta-analysis.

| Reference | No. of Sites | Attrition | Age (Average) | Age SD (±) |
| --- | --- | --- | --- | --- |
| [ Allen 2013 ] | 5 hospitals | 20% | 77.1 | 6.1 |
| [ Wong 2013 ] | *nr* | *nr* | 50.1 | 17.8 |
| [ Selinger 2013 ] | 1 hospital | 47% | 57.5 | *nr* |
| [ Sadahiro 2014 ] | 1 hospital | 5% | 66.7 | 10.7 |
| [ Ouwehand 2014 ] | 1 hospital | 11% | 50.2 | 6.5 |
| [ Georgieve 2015 ] | 1 hospital | 25% | 8.9 | 4.0 |
| [ Alberda 2018 ] | 1 hospital | 0% | 58.7 | 15.3 |
| [ Popescu 2020 ] | 1 hospital | *nr* | 65.0 | 12.0 |
| [ Wong 2021 ] | 3 hospitals | 11% | 55.0 | *nr* |
| [ Lukasik 2022 ] | 1 hospital | 10% | 2.3 | *nr* |

*No. of Sites* = Number of Sites; *Age SD* = Standard Deviation of the patient age; *nr* = not reported.

### 3.5. Patient Population

**Table 1** and **Table 2** provide breakdowns of the patient populations. Of the 10 trials, 8 were performed at one hospital (80%), while 2 (20%) were performed at multiple healthcare sites (such as specialty centres, hospitals, or specific departments) (11,18). All studies included inpatient patient analysis. The ages of the patients enrolled in the ten trials range from 3 months to 84 years of age. Most of the trials enrolled adult populations however 3 (30%) trials involved elderly patients with a mean population age ≥ 65 years of age (11,13,17). Two trials (20%) included patients from pediatric populations (15,19). The average ages of the pediatric studies were 2 and 9 years of age, respectively (15,19). The total number of patients enrolled over the 10 trials is 5,146 with 2,743 patients in the intervention group and 2,403 in the placebo groups. Nine out of the ten studies (90%) reported the participants sex categories, therefore a total of 2,368 (48%) participants were female and 2,605 (52%) participants were male. Not all clinical trials reported race or ethnicity. Seven (70%) of the trials reported duration of follow-up which ranged from 21 days to 8 weeks.

### 3.6. Antibiotic Exposure

The antibiotic exposure for the 10 trials is shown in **Table 3**. Of the ten trials, most reported the antibiotic type (n = 7, 70%) and whether single or multiple antibiotics (n = 8, 80%) were used in the patient populations. The most common antibiotic types included beta-lactam originating antibiotics: penicillin (n = 3, 30%) and cephalosporins (*n* = 4, 40%). All studies had criteria for patient selection which specified that antibiotic exposure within a certain period prior to the study start date would result in patient exclusion from the study. Of the ten studies, 50% described the most common infections that warranted antibiotic use. The most common infections include respiratory (RTI) and urinary tract infections (UTI).

**Table 3.** Summary of antibiotic exposure in trial patient populations. Includes most common antibiotic type, percentage of patient population exposed to that antibiotic, and most common infection types.

| Reference | Single or Multiple Antibiotics | Most Common Antibiotic Type | Most Common Antibiotic % | Most Common Infections Treated (if any) |
| --- | --- | --- | --- | --- |
| [ Allen 2013 ] | Multiple | Penicillins | 71.80% | respiratory, thoracic and mediastinal disorders |
| [ Wong 2013 ] | *nr* | *nr* | *nr* | *nr* |
| [ Selinger 2013 ] | Single | Penicillins | 77.29% | *nr* |
| [ Sadahiro 2014 ] | Single | Aminoglycoside and Metronidazole | 34% | surgery prophylaxis |
| [ Ouwehand 2014 ] | Single | Cephalosporins | 56.05% | respiratory, UTI, surgery prophylaxis |
| [ Georgieva 2015 ] | Multiple | Cephalosporins | 81% | respiratory, GI, ENT, UTIs |
| [ Alberda 2018 ] | Multiple | Cephalosporins | 78% | *nr* |
| [ Popescu 2020 ] | *nr* | *nr* | *nr* | *nr* |
| [ Wong 2021 ] | Multiple | *nr* | *nr* | *nr* |
| [ Lukasik 2022 ] | Multiple | Penicillins | 40% | respiratory and UTI |

*nr* = not reported.

### 3.7. Interventions

#### 3.7.1. Probiotics in CDI Prevention Trials

The full list of probiotic interventions used in the trials is outlined in **Table 4**. Out of the ten studies, fifty per cent of them (*n* = 5) used a single probiotic strain, the trials ranged from using one to eight strains of probiotics with a median of 2.5 strains. The species of probiotics used in the trials include *Bacillus, Bifidobacteria, Clostridioides, Lactobacillus, Lactocaseibacillus, Lactoplantibacillus, Ligilactobacillus, and Streptococcus*. All trials specified the strain of probiotics used in the intervention arm. Some trials (*n* = 6, 60%) used specific strains and brands of probiotics (11–13,15,16,19). Allen et al used two strains of *Lactobacillus acidophilus* CUL60/NCIMB30157 and CUL21/NCIMB30156 plus *Bifidobacterium bifidum* CUL20/NCIMB30153 and *Bifidobacterium lactis* CUL34/NCIMB20172 (11). Selinger et al used VSL#3 which included 8 separate probiotics strains which included *Bifidobacterium breve, Bifidobacterium longum, Bifidobacterium infantis, Lactobacillus acidophilus, Lactobacillus plantarum, Lactobacillus paracasei, Lactobacillus delbrueckii subsp. bulgaricus, Streptococcus thermophilus (12)*. Sadahiro et al used a specific *Bifidobacterium bifidum* from Biofermin Pharmaceutical Co., Ltc Kobe, Japan (13). Georgieva et al used a specific *Lactobacillus* strain known as *Lactobacillus reuteri* DSM 17938 from Stockholm, Sweden (15). Alberda et al used Danactive® which uses a *Lactobacillus casei* sp. Paracasei CNCM I-1518 (formally DN-114 001) (16). Lastly, Lukasik et al used an Ecologic AAD 612 preparation of antibiotic which includes 8 strains of probiotics such as *Bifidobacterium bifidum, B. lactis, Lactobacillus acidophilus, L. acidophilus, Lactocaseibacillus paracasei, Lactoplantibacillus plantarum, Lactocaseibacillus rhamnosus, and Ligilactobacillus salivarius (19)*.

**Table 4.**
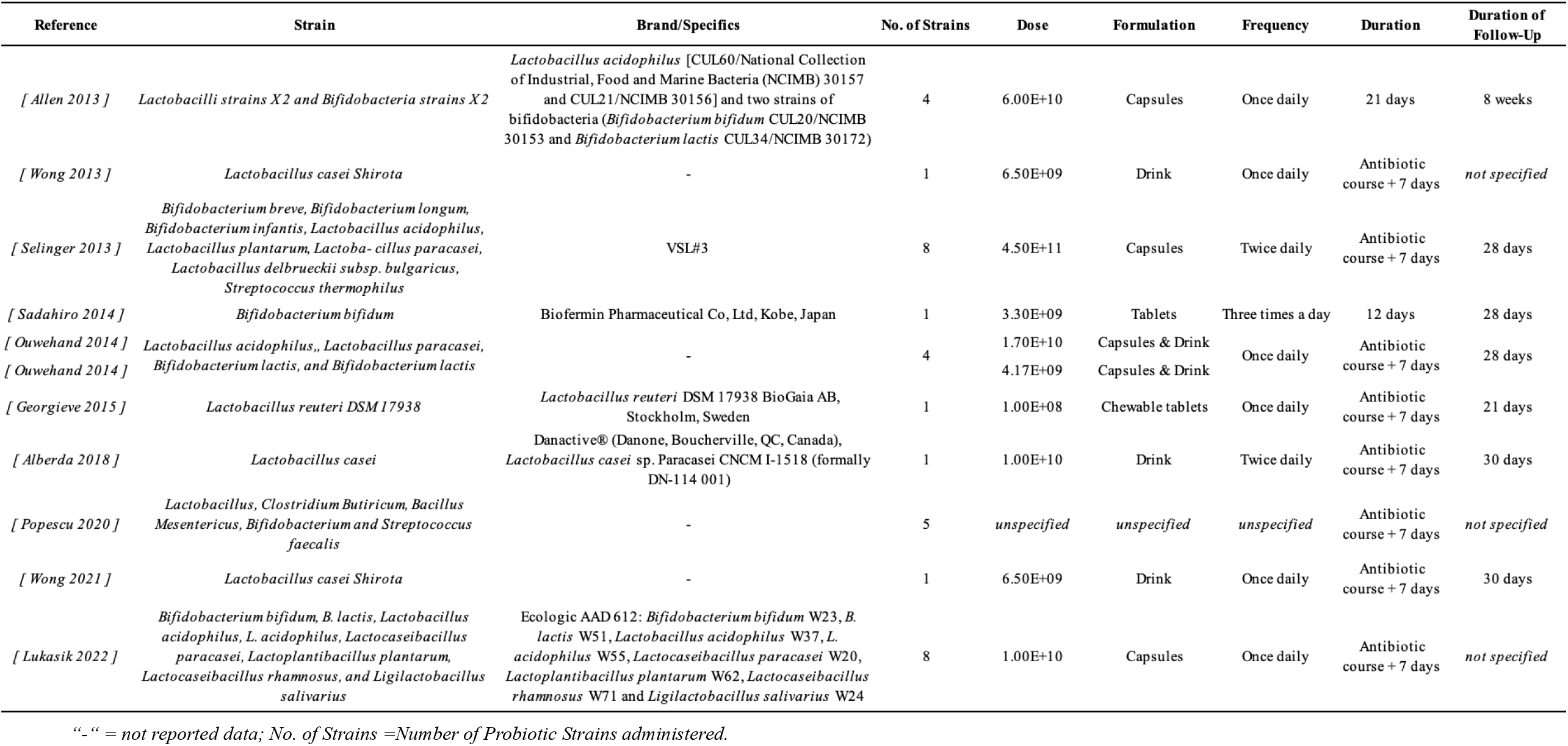
Description of probiotics administered during the treatment arms in all 10 trials. Data included is strain, brand names or other specifics, number of probiotic strains use, dosage, formulation, frequency of administration, duration of probiotic course, and duration of follow-up.

Most studies included the dose, frequency, duration of probiotic use, and formulation of probiotic all of which is described in **Table 4**. Frequency of probiotic administration ranged from once to three times daily. Formulation of probiotics varied between capsules, chewables, drink, or tablets.

One study did not mention the formulation of probiotic used (17). A large proportion of studies used either capsules (*n* = 4, 40%) or drink (*n* = 4, 40%).Most of the trials (*n =* 8, 80%) administered probiotics for the duration of the antibiotic course + 7 days post-antibiotic course. One study administered probiotics for 21 days regardless of antibiotic used (11). Another study administered probiotics for a total of 12 days (pre-operatively and post-operatively) for participants’ elective colorectal surgeries (13). The probiotic dosage range varied from 1.00E+08 colony forming unites (CFUs) to 4.50E+11 CFUs. Only one trial did not specify the dosage of probiotic used as an intervention (17). Duration of follow-up varied from 21 days to 8 weeks, three studies, all of which were abstracts, did not specify the duration of follow-up (10,17,19). None of the trials reported negative safety outcomes with probiotics as an intervention treatment.

For *Clostridioides difficile* outcomes, all studies mentioned *C. difficile* testing and confirmation of stool samples for CDI.

### 3.8. Pooled Efficacy of Probiotics for CDI Prevention

#### 3.8.1. Meta-analysis

A meta-analysis of the 10 trials of probiotics *versus* controls was performed and the pooled results showed a high degree of heterogeneity (I^2^ = 99.94%, *p* < 0.0001), so a random-effects model (RE model) was used. The funnel plot analysis, which also used the random-effects model, shows that there are several smaller trials with low standards errors and small effect sizes, while there is a single dot that shows a substantial effect size with a large standard error (**Figure 2**). This dot is an outlier and could have a significant impact on the overall results of the meta-analysis. This dot was removed from the data and meta-analysis was performed again (17). Arguments for the removal of this data include the author(s) did not provide sufficient data within the abstract for proper meta-analysis while also weakening the results of the meta-analysis. The degree of heterogeneity dropped, however it remains high, so the random-effects model is still used (I^2^ = 95.85%, *p <* 0.0001).

**Figure 2.**
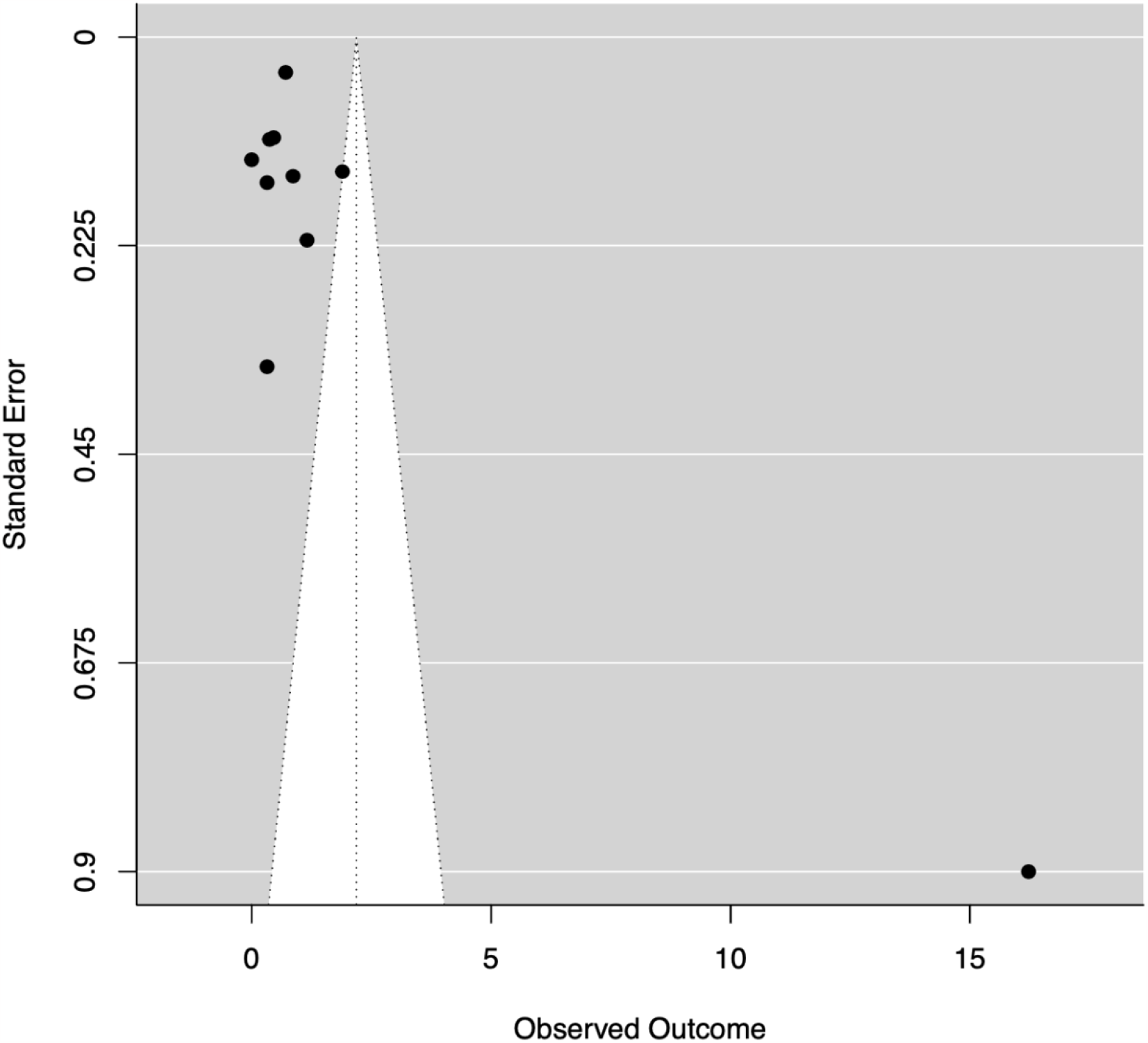
Funnel plot analysis using the random-effect analysis which includes data from all 10 trials included in the meta-analysis. Observed outcome is relative risk.

Overall, the data suggests that probiotics have a positive effect in preventing CDI during the administration of antibiotics, but the strength of this association varies across the trials (*estimated effect* of 0.68, *95% CI* (0.31-1.06)). Some studies show a stronger effect, while others show no significant effect. This can be seen in the Forest Plot (random-effects model) in **Figure 3**.

**Figure 3.**
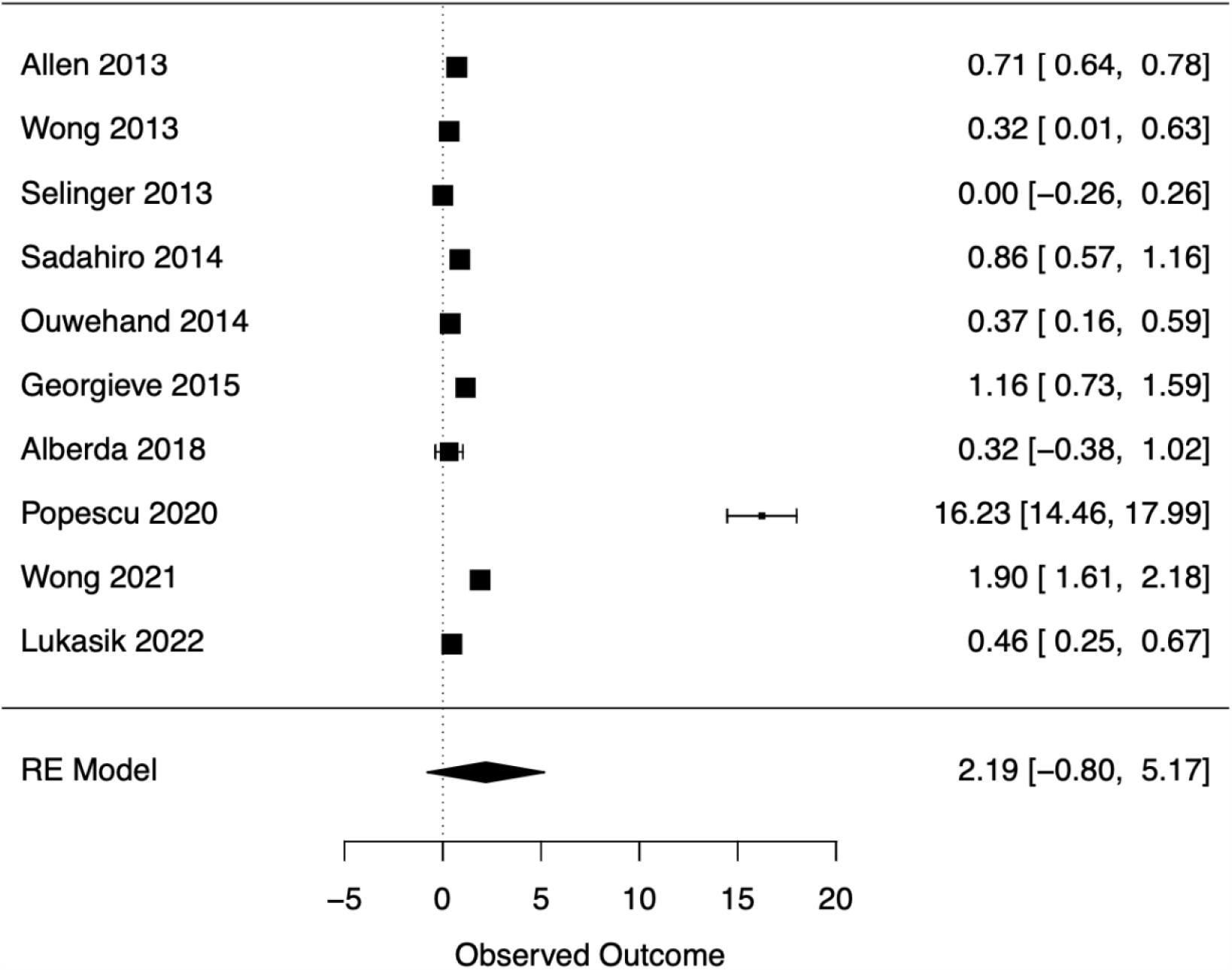
Forest Plot using the random-effects model used to analyze the total observed outcome across all 10 trials. RE Model generated an observed outcome (relative risk) of 2.19 with a 95% confidence interval of -0.80 to 5.17.

A new Funnel Plot analysis and Forest Plot (both using random-effects model) was generated and can be seen in **Figure 4A** and **4B**. A Rank Correlation Test for Funnel Plot Asymmetry was performed and generated Kendall’s *tau* = 0.0556, *p* = 0.9195, meaning that there is weak and non-significant evidence of asymmetry within the funnel plot. The trials included in the analysis are distributed fairly round the combined effect estimate. There is no indication of publication bias that could be affecting the meta-analysis (9). In addition, the Fail-safe N Calculation using the Rosenthal Approach was calculated (N: 1042), which shows that a large number of nonsignificant studies will not influence the meta-analysis too much (20).

**Figure 4A and 4B.**
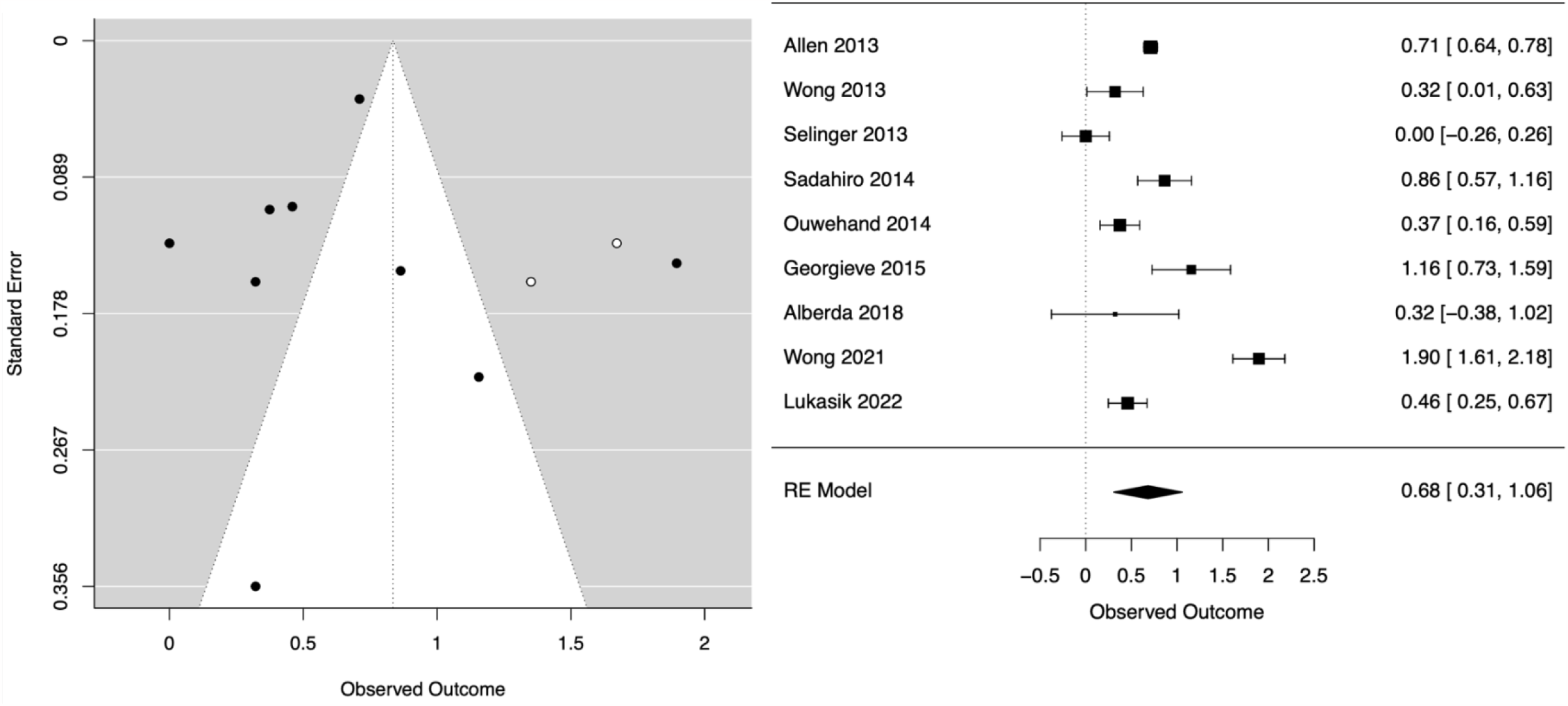
Figure 4A (left) is a repeat Funnel Plot analysis without the data from Popescu et al trial (outlier) and Figure 4B (right) is a repeat Forest Plot analysis without the data from the Popescu et al trial (outlier). Observed outcome is relative risk as reported in the trials. Compare to **Figure 2** and **3**.

#### 3.8.2. Sub-group analysis

Due to the high level of heterogeneity, sub-group analysis was performed. Meta-regression analysis results for the use of probiotics in the prevention of CDI during antibiotic administration did not find *any significant associations* between the study population type (adult *versus* pediatric, *p* = 0.2209) or size of probiotic dose (≥5.68E+10 *versus* <5.68E+10, *p* = 0.3583. Sub-group analysis of numbers of probiotic strains showed that both using one (estimated effect size of 0.9248, CI (95%): 0.1285-1.7211, *p* = 0.0228) and two or more probiotic strains (estimated effect size of 2.1366, *p* = 0.0000) are effective in preventing CDI, but using multiple strains appears to have a stronger impact. The variance of 0.1755 represents a level of uncertainty that could mean that the effect is slightly lower or slightly higher. Both studies show high levels of heterogeneity, with an overall result of 98%. This could be attributed to the fact that *different* probiotic strains were used across the board for all the trails (see **Table 4** for details on probiotics used). The level of heterogeneity in these trials could be due to the variation in probiotic strains, the length of time probiotics was administered, the dose of the intervention arm, different antibiotics administered, the sample populations in terms of infection types and age, and frequency of probiotic administration.

## 4. Discussion

*Clostridioides difficile* infections continue to be a significant concern in hospital settings, causing nosocomial outbreaks and posing considerable healthcare burdens across the globe. The Gram-positive, spore-forming, anaerobic bacteria are responsible for most cases of antibiotic-associated diarrhea. Antibiotics disrupt the normal balance of the gut microbiota, allowing *C. difficile* to proliferate and cause clinical symptoms, especially in vulnerable populations. CDI is associated with symptoms such as diarrhea, abdominal pain, fever, nausea, and anorexia, and in severe cases, it can lead to intensive care unit admission, colectomy surgery, or death. The elderly, individuals on broad-spectrum antibiotics, and those with reduced gastric acidity are particularly susceptible to CDI. The increasing incidence of CDI highlights the need for effective prevention strategies in healthcare settings. Standard preventative practices include infection control, antibiotic stewardship, and proper hand hygiene. Despite these measures, *C. difficile* outbreaks still occur, emphasizing the importance of exploring novel methods in prevention.

Probiotics, defined as “good bacteria” that naturally reside in the human gut and contribute to the maintenance of a healthy microbiota, have been proposed as a potential preventive measure against *C. difficile* infections. Probiotics have long been questioned for their efficacy in preventing CDI and AAD, but their use in clinical practice remains controversial. This meta-analysis aimed to provide further insights into the role of probiotics in the prevention of CDI. The meta-analysis pooled data from ten randomized controlled trials, which included a total of 5,146 participants. The probiotic strains tested in these trials primarily included *Lactobacillus spp*. and *Bifidobacterium spp*. The meta-analysis revealed a pooled effect size of 0.68 (95% CI: 0.31-1.06), indicating a positive effect of probiotics in preventing CDI during antibiotic treatment. However, the results demonstrated significant heterogeneity among the trials, indicating that the effect of probiotics on CDI prevention varied across the trials. The observed heterogeneity in the meta-analysis could be attributed to several factors, such as the variation in probiotic strains, dosages and formulations, duration of probiotic administration, and the diversity of antibiotic regimens and patient populations among the studies. To better understand the influence of these factors on probiotic efficacy, sub-group analyses were conducted. The sub-group analysis of probiotic strains revealed that both single-strain and multi-strain probiotics were effective in preventing CDI, with multi-strain formulations showing a stronger impact. However, it is essential to interpret these findings with caution, as different probiotic strains were used across the trials, and the overall quality of evidence remains uncertain.

The findings of this meta-analysis suggest that probiotics may possibly be beneficial in preventing CDI during antibiotic administration. However, due to the significant heterogeneity among the included studies, additional research is needed to establish standardized protocols for probiotic use in CDI prevention. Future studies should aim to address the variability in probiotic strains, dosages, and formulations to identify the most effective approach. Moreover, large-scale, well-designed clinical trials with consistent probiotic interventions and robust outcome measures are warranted to provide more definitive evidence. Additionally, a focus of future research should determine the possible interactions between probiotics and specific antibiotics to determine optimal combinations that maximize preventive efficacy for antibiotic-associated diarrhea and CDI.

Despite these valuable insights, this meta-analysis has some limitations. First, the considerable heterogeneity among the trials undermines the strength of the overall findings. Second, the number of studies included in the analysis was relatively small, limiting the generalizability of the results. Third, some of the included trials were abstracts, which may lack comprehensive data and rigorous reporting. Lastly, the quality of the trials and potential biases were not systematically assessed, potentially affecting the overall validity of the results.

## 5. Conclusions

*Clostridioides difficile* infections (CDI) remain a significant concern in healthcare settings, necessitating effective and novel prevention strategies. Probiotics have been investigated as a potential preventive measure in several random-controlled trials. The meta-analysis performed herein concludes and suggests that probiotics may have a positive effect in preventing CDI during antibiotic treatment. However, the observed heterogeneity in the results highlights the need for further research to establish standardized probiotic protocols and to address the variability in probiotic strains, dosages, and formulations. Large-scale, well-designed clinical trials are essential to provide more definitive evidence on the efficacy of probiotics in CDI prevention, enabling healthcare professionals to implement evidence-based intervention strategies to mitigate the impact of this nosocomial infection.

## Data Availability

All data produced in the present study are available upon reasonable request to the authors.

